# Testing, Tracing, and Vaccination Targets for Containment of the US Monkeypox Outbreak: A Modeling Study

**DOI:** 10.1101/2022.08.01.22278199

**Authors:** Melanie H. Chitwood, Jiye Kwon, Alexandra Savinkina, Jo Walker, Alyssa Bilinski, Gregg Gonsalves

## Abstract

We estimate the levels of community testing, contact tracing, and vaccination required to reduce the effective reproduction number (Rt) of Monkeypox Virus (MPXV) below 1 for high risk men who have sex with men. We found that the critical threshold to vaccinate depends on the basic reproduction number of MPXV and on other public health measures. This analysis provides a framework for quantitative targets toward MPXV containment.

## Background

As of July 31 2022, there have been over 5,000 confirmed cases of monkeypox across 48 U.S. states, predominantly among men who have sex with men (MSM)^1^. Reducing transmission of monkeypox virus (MPXV) is crucial to protect the health of the MSM population and reduce the risk of transmission in the general US population. Public health measures – testing, contact tracing, vaccination – can slow the spread of MPXV and officials have called for rapid scale-up of these efforts to support containment^2^. However, target levels of these measures have not been established.

## Objective

We estimate the levels of community testing, contact tracing, and vaccination required to reduce the effective reproduction number (R_t_) of MPXV below 1 for high risk MSM (HR-MSM).

## Methods and Findings

We adapted a deterministic branching model^3^ to describe the transmission of MPXV in HR-MSM (Supplement). Infectious individuals are detected with a probability that varies based on community detection rate and whether their source case underwent contact tracing (Figure 1). We assume that community detection of a case reduces the number of secondary infections by 50%, identifying a case early through contact tracing reduces the number of secondary infections by 90%, and all detected cases have the same probability of having their contacts traced. We used Byrd optimization to solve for the level of vaccine coverage (assuming 100% efficacy) for HR-MSM that would reduce R_t_ below 1 at different estimates of the basic reproduction number (R_0_) in HR-MSM^4^ and rates of contact tracing and community detection. Previous work indicates that R_0_ remains below 1 in the general population; to reflect uncertainty in HR-MSM, we vary R_0_ from 1.2-2.0.

**Figure 1.**
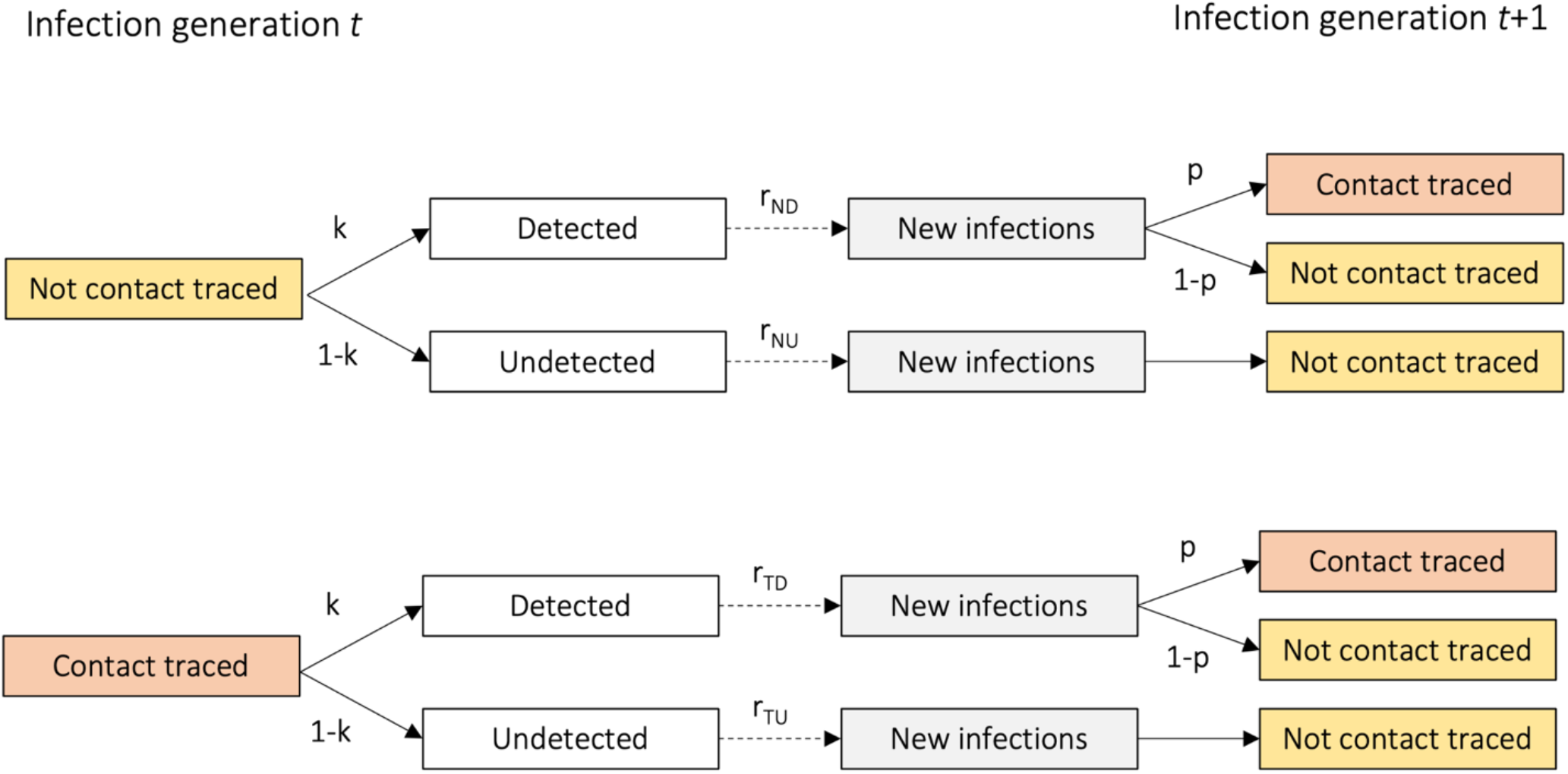
Model structure.

We estimate there are 498,000 HR-MSM in the US (Supplement). We found that testing and contact tracing without vaccination can reduce R_t_ below 1 in this population if R_0_ ≤ 1.4, at least 40% of cases are detected through community testing, and at least 50% of case contacts are traced. In the absence of a robust public health response (10% community detection; no contact tracing), we estimate that the critical threshold to vaccinate ranges from 12% (R_0_ = 1.2) to 47% (R_0_ = 2.0). With a moderate response, (at least 20% community detection;at least 25% of cases contact traced) we estimate the critical threshold to vaccinate ranges from 5% (R_0_ = 1.2) to 43% (R_0_ = 2) of the HR-MSM population (Figure 2). In this scenario, 49,800 – 428,080 doses of the Jynneos vaccine would need to be administered to 24,900 – 214,040 HR-MSM.

**Figure 2.**
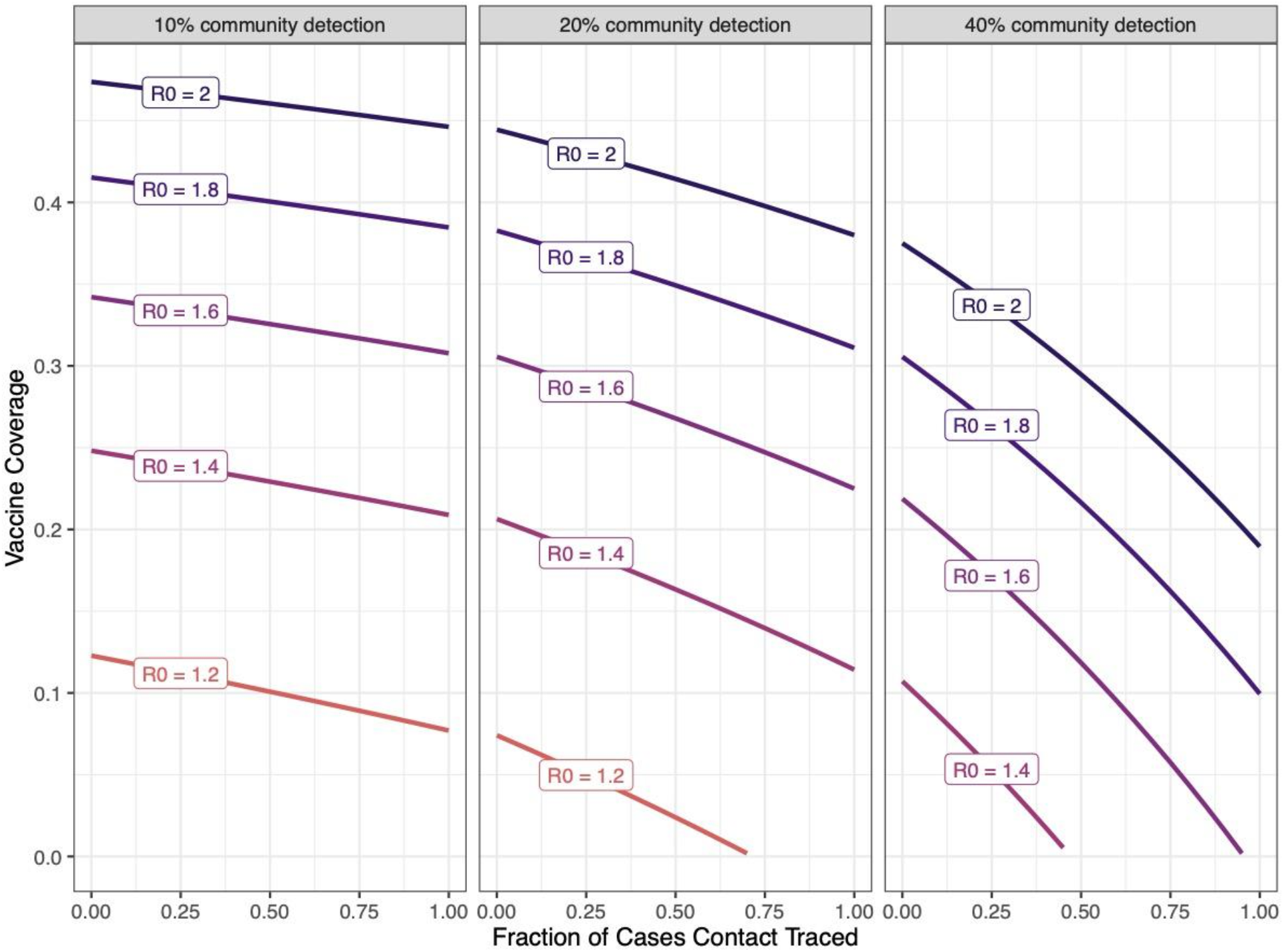
Frontier of Contact Tracing, Community Testing, and Vaccination Coverage to Reduce the Effective Reproduction Number Below 1. The x-axis displays the fraction of cases traced and the y-axis vaccine coverage. The community detection rate is varied across facets. Each line shows the efficient frontier for a given value of R_0_. Each point on the line can be interpreted to mean the fraction of contacts traced and the vaccination rate at which Rt remains below 1.

The critical threshold to vaccinate was not sensitive to assumptions about the fraction of secondary cases detected through contact tracing (Figure S1), but was sensitive to assumptions about how much less infectious detected cases are relative to undetected infections. With 20% community detection, 25% of case contacts traced, and R_0_ = 1.6, a 25% reduction in secondary infections suggests a 32% threshold, while a 75% reduction suggests a 25%threshold (Figure S2).

## Discussion

We project the impact of public health measures to reduce the spread of MPXV in the US. We found that the critical threshold to vaccinate depends on the basic reproduction number of MPXV and on other public health measures. Nevertheless, rapid distribution of vaccinations to at least one third of the HR-MSM (at least 329,000 doses) with increased testing and contact tracing, can support containment efforts in most modeled scenarios. As of July 29, the US government has shipped 336,710 doses, although the number of administered doses and the specific profile of recipients are unclear^5^.

This analysis has a number of limitations. We estimate vaccine needs based on the assumption that spread outside the HR-MSM population is negligible. If there is sustained transmission of MPXV in other populations at risk (e.g. low-risk MSM; in congregate settings), increased vaccination efforts may be required. We further assume that vaccines have 100% efficacy and are efficiently targeted to HR-MSM. Last, key parameters remain uncertain, although we illustrate sensitivity of our results to variations in these, and can adjust estimates as more information becomes available. As a result, we believe this analysis provides a useful framework for quantitative targets toward MPXV containment.

## Data Availability

All data produced in the present work are contained in the manuscript.

## Supplement

Table S1. Model parameter values

Table S2. Estimation of secondary infections.

Figure S1. Vaccination and contact tracing efforts required to reduce Rt below 1, by R_0_ and community detection rates.

Figure S2. Vaccination and contact tracing efforts required to reduce Rt below 1 by percentage of contacts traced.

