## Supplement for "Testing, Tracing, and Vaccination Targets for Containment of the US Monkeypox Outbreak: A Modeling Study": Supplement_MPXV.docx

**Supplementary Materials**

**Methods**

To describe the transmission of monkeypox virus (MPXV) in a population of high risk men who have sex with men (HR-MSM), we adapted a deterministic branching model first developed for SARS-CoV-2^3^. The model by Bilinski et al. has compartments to account for detection at the asymptomatic, presymptomatic, or symptomatic stage. As transmission of MPXV is presumed to only occur among symptomatic individuals^6^, we removed the asymptomatic and presymptomatic states and transitions. We made additional changes to parameter values to account for the epidemiological differences between the two viruses (Table S1).

The model estimates the number of secondary infections arising from cases. Cases can be detected either through community testing programs or as a result of contact tracing. Community testing identifies individuals once they are infectious; these individuals take measures to reduce their contacts by 50%. Contact tracing identifies individuals prior to the onset of infectiousness; these individuals enter quarantine and reduce their contacts by 90%. Undetected cases do not reduce their contacts and do not undergo contact tracing; the secondary infections arising from undetected cases can be detected through community testing only. All detected cases have the same probability of having their contacts traced.

We estimated the rate of vaccination and contact tracing needed to lower R_t_ below 1, assuming community detection rates of 10%, 20%, and 40%. We performed sensitivity analysis on the fraction of cases identified through contact tracing and the reduction in secondary infections generated by detected cases (Figures S1 and S2).

We estimate the number of individuals that need to be vaccinated to support containment of MPXV in the HR-MSM population. To estimate the size of this population, we used nationally representative survey data suggesting that MSM make up 3.9% of the adult male population in the US^7^. Multiplying this percentage by the 2021 population estimates of all US adult males (n = 127,808,000) from the US Census Bureau^8^, we estimated that there are 4.98 million adult MSM in the US. Next, we considered polling data that suggest 10% of MSM in the US have 100 or more lifetime sexual partners^9^; we used this to define ‘high risk’ and estimated that there are 498,000 HR-MSM in the US.

Finally, we estimate the number of vaccine doses needed to support containment. Because the Jynneos vaccine requires two doses given at least four weeks apart^10^, the number of doses needed is twice the number of individuals that need to be vaccinated. We make the simplifying assumptions that the Jynneos vaccine is 100% efficacious against infection, that protection is conferred immediately following the first dose, that no doses are wasted, and that doses are only given to HR-MSM. As a result, our estimates represent the lowest number of doses needed in each scenario.

**Table S1. Model parameter values**

| **Parameter** | **Value(s)** | **Source** |
| --- | --- | --- |
| Fraction of cases detected (k) | 10%, 20%, 40% | Assumed |
| R0 | 1.2, 1.4, 1.6, 1.8, 2.0 | (4) |
| Fraction of cases successfully traced (p) | 50% | Assumed |
| Duration of infectiousness (d) | 21 days | (11) |
| Relative number of secondary infections  from detected infections compared to  undetected infections (q)   - note: this applies to cases that aren’t contact traced but are detected through testing, as well as cases that are contact traced (all of which we assume are detected, see above). As a result, the overall transmissibility ratio $\frac{R_{contact traced}}{R_{not traced or otherwise detected}}$ is computed as the product of this parameter and ($1-\epsilon$), see below | 0.5 | Assumed |
| Average daily rate of transmission for  symptomatic cases not traced (b) | [calibrated](https://cdn.jamanetwork.com/ama/content_public/journal/jamanetworkopen/938527/zld200145supp1_prod_1602622418.91358.pdf?Expires=1659385848&Signature=atCMeCrb3NTdHmTU3WuaPAq2Xot3ipPmQOtVZs6UhfKj5X10i1Jv1fY4VqP39vgkn~EbrAaF~BDJKFxDMSRL0LL7jV59IuFSQZqRBF1o--uW3BPSooxomxmfIar3zv299EtzaC1G6DtgMHr2jhHCmZgUBZuKgqOcPC4SSkS8i1VEz9XEnNYP2UOACi99uVOxYc4tu6gQvXF1Fk-YMIBLJpFE6JvL~xBlE5yNcdwlwdgxntTJTm3804jbiSVYJXoMz~RitXHfPwix15NZYQM4xsP~WgNONu3loEeLrtbsBfyHWVeABiKqlfGgybDQzJOklI0-35re699KdZeYG~PiFA__&Key-Pair-Id=APKAIE5G5CRDK6RD3PGA) |  |
| Isolation and quarantine efficacy ($\epsilon$, only applicable to cases that are contact traced) | 90% | Assumed |
| Vaccination % (v) | Varies |  |

**Table S2. Estimation of secondary infections.**

| Category | Formula |
| --- | --- |
| Not contact traced, detected | rND = (1-v)bdq |
| Not contact traced, undetected | rNU = (1-v)bd |
| Contact traced, detected | rTD = (1-e)(1-v)bdq |
| Contract traced, undetected | rTU = (1-e)(1-v)bd |

Where:

b= Average daily rate of transmission for cases not traced

d=duration of infectiousness

q=Relative number of secondary infections from detected infections compared to undetected infections

e= Isolation and quarantine efficacy

**Figure S1. Vaccination and contact tracing efforts required to reduce Rt below 1 by percentage of contacts traced.** Assuming 20% community detection and R_0_ = 1.6. The x-axis displays the fraction of cases traced and the y-axis vaccine coverage. Each line shows the efficient frontier for a given value of the probability that an infected traced contact is detected.

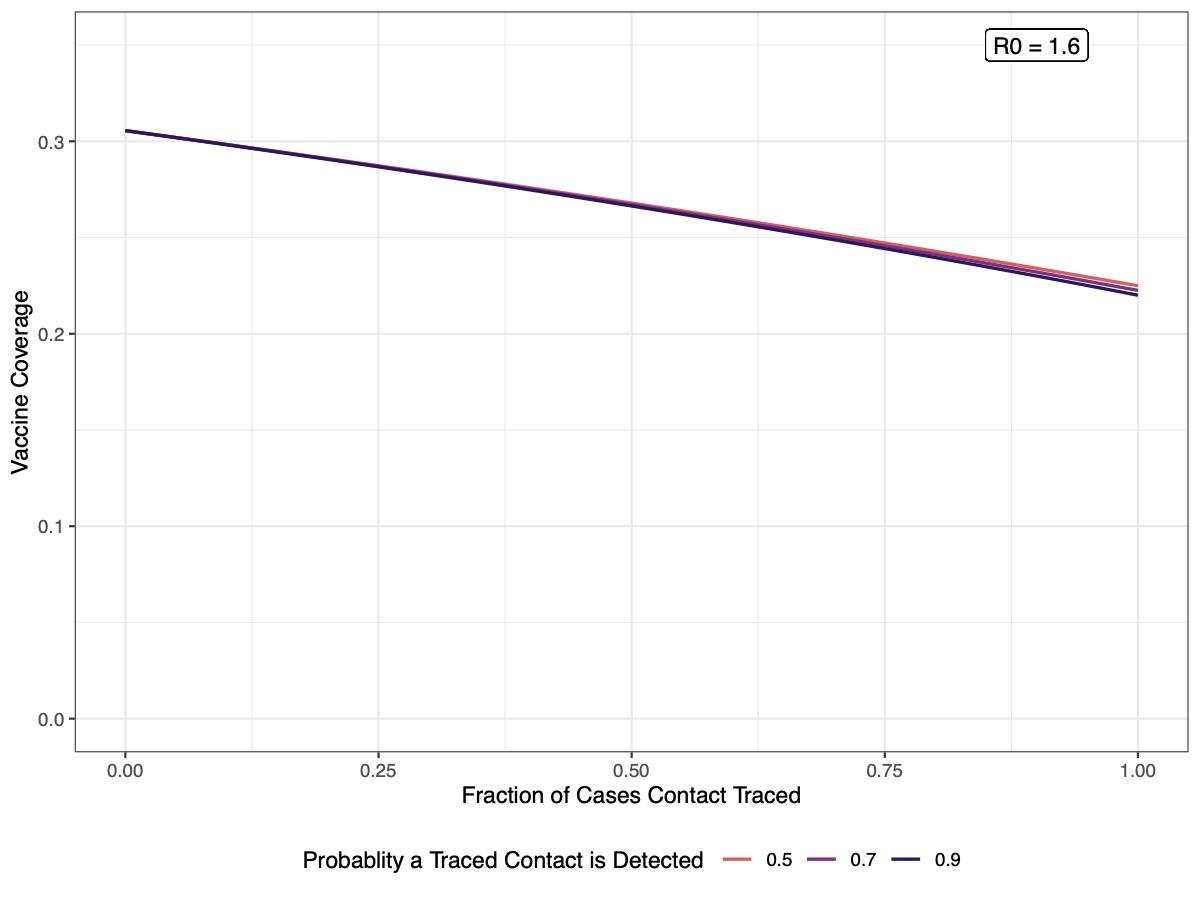

**Figure S2. Vaccination and contact tracing efforts required to reduce Rt below 1 by reduction in secondary infections due to detection.** Assuming 20% community detection, 25% of case contacts traced, and R_0_ = 1.6. The x-axis displays the fraction of cases traced and the y-axis vaccine coverage. Each line shows the efficient frontier for a given value of the reduction in secondary infections among detected cases.

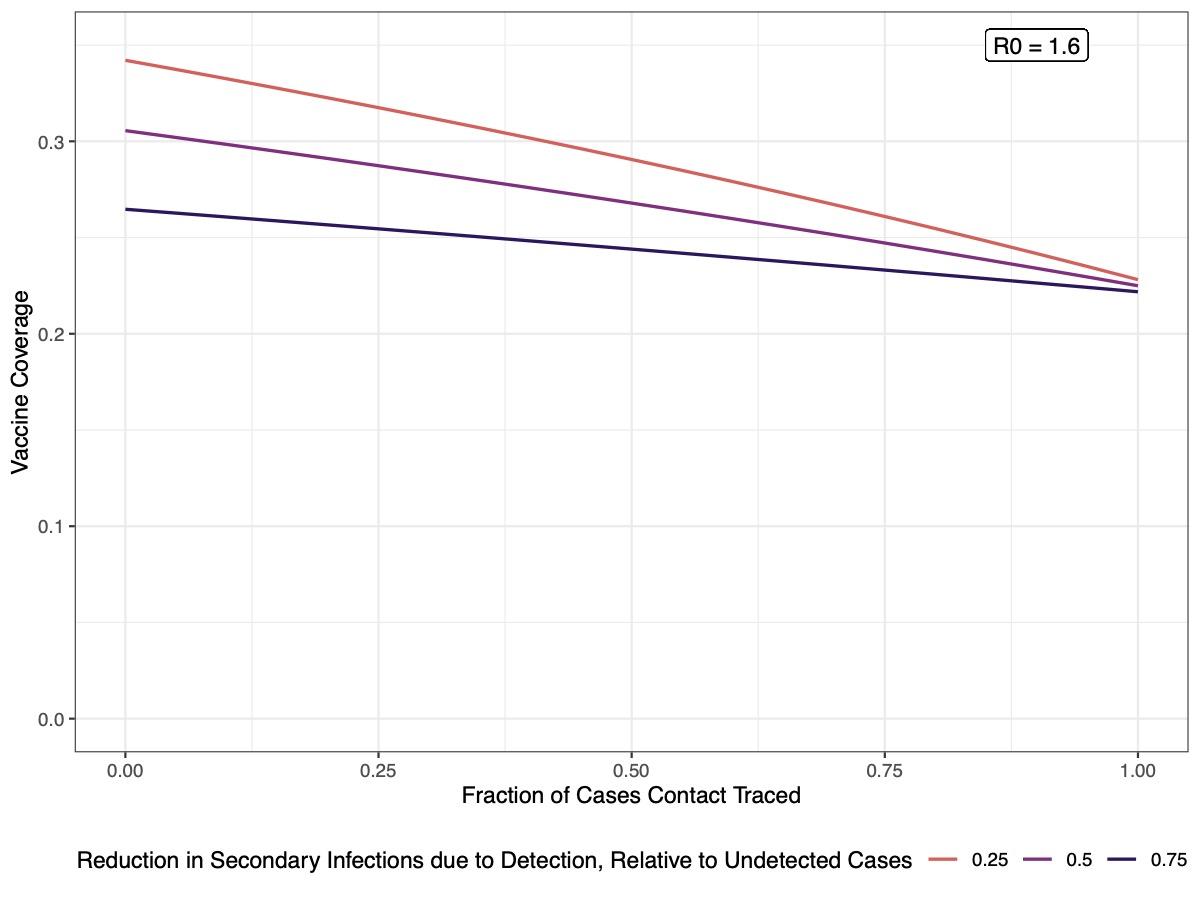
